# High Rate of Asymptomatic Carriage Associated with Variant Strain Omicron

**DOI:** 10.1101/2021.12.20.21268130

**Authors:** Nigel Garrett, Asa Tapley, Jessica Andriesen, Ishen Seocharan, Leigh H. Fisher, Lisa Bunts, Nicole Espy, Carole L. Wallis, April Kaur Randhawa, Nzeera Ketter, Margaret Yacovone, Ameena Goga, Linda-Gail Bekker, Glenda E. Gray, Lawrence Corey

## Abstract

The early widespread dissemination of Omicron indicates the urgent need to better understand the transmission dynamics of this variant, including asymptomatic spread among immunocompetent and immunosuppressed populations. In early December 2021, the Ubuntu clinical trial, designed to evaluate efficacy of the mRNA-1273 vaccine (Moderna) among persons living with HIV (PLWH), began enrolling participants. Nasal swabs are routinely obtained at the initial vaccination visit, which requires participants to be clinically well to receive their initial jab. Of the initial 230 participants enrolled between December 2 and December 17, 2021, 71 (31%) were PCR positive for SARS-CoV-2: all of whom were subsequently confirmed by S gene dropout to be Omicron; 48% of the tested samples had cycle threshold (CT) values <25 and 18% less than 20, indicative of high titers of asymptomatic shedding. Asymptomatic carriage rates were similar in SARS-CoV-2 seropositive and seronegative persons (27% respectively). These data are in stark contrast to COVID-19 vaccine studies conducted pre-Omicron, where the SARS-CoV-2 PCR positivity rate at the first vaccination visit ranged from <1%-2.4%, including a cohort of over 1,200 PLWH largely enrolled in South Africa during the Beta outbreak. We also evaluated asymptomatic carriage in a sub study of the Sisonke vaccine trial conducted in South African health care workers, which indicated 2.6% asymptomatic carriage during the Beta and Delta outbreaks and subsequently rose to 16% in both PLWH and PHLWH during the Omicron period.

These findings strongly suggest that Omicron has a much higher rate of asymptomatic carriage than other VOC and this high prevalence of asymptomatic infection is likely a major factor in the widespread, rapid dissemination of the variant globally, even among populations with high prior rates of SARS-COV-2 infection.

---

The emergence of the B.1.1.529 (Omicron) SARS-CoV-2 variant, first identified in Botswana and South Africa^1^ and as of Dec 18, 2021, found in over 89 countries, has raised concern for a new global wave of COVID-19. The large number of deletions and mutations, some of which overlap with those seen in prior Variants of Concern (VoC), are worrisome for potentially increased transmissibility, viral binding affinity, and immunologic escape^2,3^. These concerns have intensified given that South Africa is experiencing a new rise in COVID-19 cases at a rate faster than any of the three previous waves^4^, and despite ongoing mask mandates and high antibody seropositivity due to prior infection or vaccination^5,6^. The early widespread dissemination of Omicron including rapid spread in Europe and the United States (US) indicates there is an urgent need to better understand the transmission dynamics of Omicron, including asymptomatic spread among immunocompetent and immunosuppressed populations.

On December 2, 2021 we began enrollment into the Ubuntu multicenter Phase 3 clinical trial in sub-Saharan Africa (with all sites initially enrolling located in South Africa) to assess the relative efficacy of the COVID-19 mRNA vaccine mRNA-1273 (Moderna) in persons (adults) living with HIV (PLWH) and/or with at least one comorbidity known to be associated with severe COVID-19. A smaller population of HIV negative persons is also part of the trial. Previously vaccinated patients are excluded. Baseline testing includes HIV screening, CD4^+^ T-cell count and HIV viral load (if HIV positive), and collection of a nasal swab for reverse-transcriptase polymerase chain reaction (RT-PCR) testing. The Assure Ecotest IgG/IgM Rapid Test (Assure Tech) was used to determine baseline SARS-CoV-2 antibody status. Study participants must be clinically well and have no signs/symptoms of COVID-19 to be vaccinated upon enrollment.

As of December 17, 2021, a total of 330 adults were enrolled across seven South African provinces. The median age of participants was 39 years (range 18-76 years); 79% were female sex assigned at birth. Baseline nasal swab data were available for 230/330 enrolled participants in five provinces. Overall, 31% (71) of participants had evidence of acute SARS-CoV-2 infection by RT-PCR (**Table 1a**), with the highest percentage in the Gauteng province. Detection of SARS-CoV-2 was similar among those who were seropositive vs seronegative to SARS-CoV-2, and no relationship between detection and CD4+ T-cell count was noted (**Table S1a**). Of the identified infections, 62 were subjected to additional testing to evaluate S gene dropout; 56 samples were successfully amplified for the Orf and N genes by TaqPath(tm) COVID 19 CE IVD RT PCR (ThermoFisher); all had S gene dropout, suggestive of Omicron infection^7^. The median RT-PCR cycle threshold (Ct) value was 25·8 (range 14·4 – 34·9), with Ct values <25 in 48% and ≤20 in 18% of participants **(Figure S1)**.

**Table 1a.** SARS-CoV-2 PCR positivity by South African province and participant serostatus.

|  | Total participants | SARS-CoV-2 PCR+ | Prevalence |
| --- | --- | --- | --- |
| **Serostatus** |  |  |  |
| SARS-CoV-2 Seronegative | 144 | 44 | 31% |
| SARS-CoV-2 Seropositive | 86 | 27 | 31% |
| **Total** | **230** | **71** | **31%** |
| **Province** |  |  |  |
| Gauteng | 53 | 31 | 58% |
| KwaZulu Natal | 111 | 22 | 20% |
| Eastern Cape | 17 | 7 | 41% |
| Western Cape | 48 | 11 | 23% |
| Mpumalanga | 1 | 0 | 0% |
| **Total** | **230** | **71** | **31%** |

Nasal swab sampling at the initial vaccination visit has been used in several COVID-19 vaccine efficacy trials to define persons infected at the time of study entry^8-12^. Studies conducted before Omicron typically exhibited asymptomatic carriage of pre-Omicron variants in ≤1% of participants^8,10,12^ (**Table 1b**), including a 1,227 PLWH subgroup in the Ensemble 1 study, largely enrolled during the Beta outbreak in South Africa. In addition to these CoVPN studies, the Sisonke study^11^ conducted exclusively in South Africa between June and August 2021 during the Delta outbreak demonstrated an asymptomatic carriage rate of 2·4% in the subgroup sampled on day of vaccination. To date, of the 577 participants of the Sisonke subgroup being resampled from mid-November to Dec 7, 2021 (time of Omicron outbreak) at the 6-month follow up visit, 91 (16%) of the 577 participants seen had SARS-COV-2 detected in their nasal swab sample (**Table S2**). The frequency of PCR positivity with Omicron was similar between PLWH (27 of 169: 16%) and HIV negative participants (62 of 405: 15·3%).

**Table 1b.**
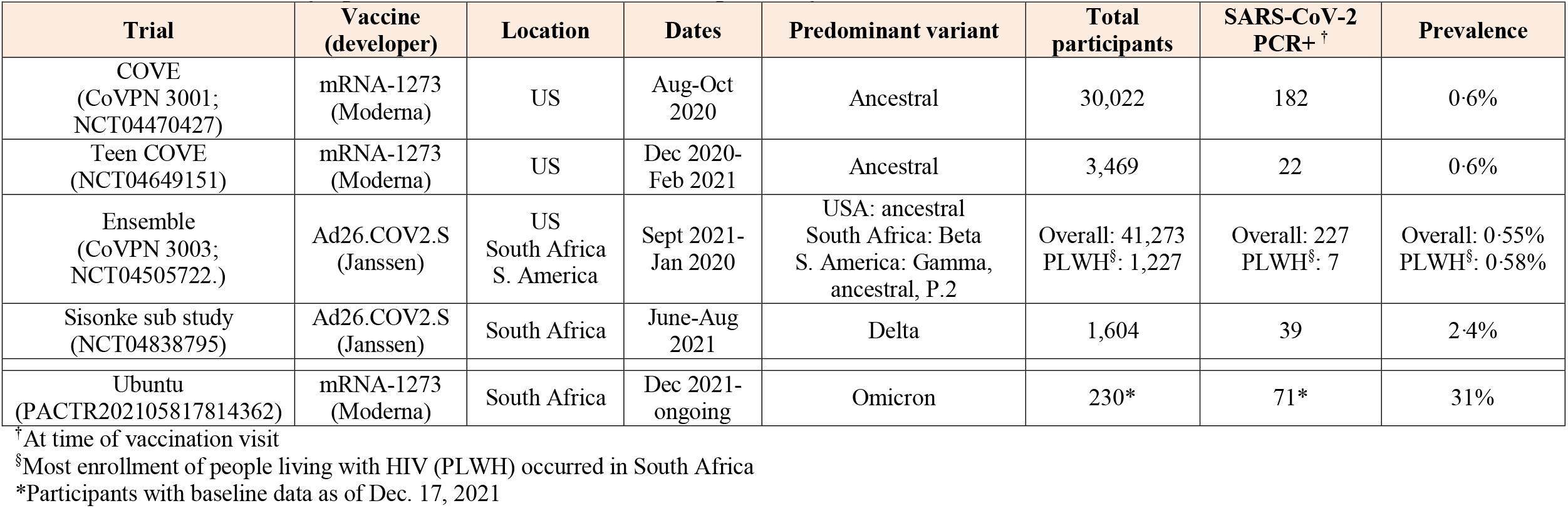
Prevalence of asymptomatic SARS-CoV-2 infection upon entry in vaccine studies.

| Trial | Vaccine (developer) | Location | Dates | Predominant variant | Total participants | SARS-CoV-2 PCR+<sup>†</sup> | Prevalence |
| --- | --- | --- | --- | --- | --- | --- | --- |
| COVE (CoVNP 3001; NCT04470427) | mRNA-1273 (Moderna) | US | Aug-Oct 2020 | Ancestral | 30,022 | 182 | 0.6% |
| Teen COVE (NCT04649151) | mRNA-1273 (Moderna) | US | Dec 2020-Feb 2021 | Ancestral | 3,469 | 22 | 0.6% |
| Ensemble (CoVNP 3003; NCT04505722.) | Ad26.COV2.S (Janssen) | US<br>South Africa<br>S. America | Sept 2021-Jan 2020 | USA: ancestral<br>South Africa: Beta<br>S. America: Gamma, ancestral, P.2 | Overall: 41,273<br>PLWH<sup>§</sup>: 1,227 | Overall: 227<br>PLWH<sup>§</sup>: 7 | Overall: 0.55%<br>PLWH<sup>§</sup>: 0.58% |
| Sisonke sub study (NCT04838795) | Ad26.COV2.S (Janssen) | South Africa | June-Aug 2021 | Delta | 1,604 | 39 | 2.4% |
| Ubuntu (PACTR202105817814362) | mRNA-1273 (Moderna) | South Africa | Dec 2021-ongoing | Omicron | 230* | 71* | 31% |

<sup>†</sup>At time of vaccination visit<sup>§</sup>Most enrollment of people living with HIV (PLWH) occurred in South Africa
\*Participants with baseline data as of Dec. 17, 2021

These findings strongly suggest that Omicron has a much higher rate of asymptomatic carriage than other VoC and this high prevalence of asymptomatic infection is likely a major factor in the widespread, rapid dissemination of the variant globally, even among populations with high prior rates of SARS-COV-2 infection. Many of these asymptomatic carriers have high nasal viral titers; suggesting that subclinical carriage could be a major factor in the rapid spread of Omicron globally.

The effects of vaccination on the prevalence or titers of asymptomatic infection are still unknown. All the samples from the Ubuntu study were from non vaccinated persons and the data from the Sisonke study are only a small subset (<1% of the total study) and hence too small to calculate any estimate of vaccine effectiveness. Obtaining data on asymptomatic carriage and transmissibility among vaccinated persons is urgently needed. The importance of non-pharmaceutical interventions and the installation of rapid detection strategies for such carriage in high-risk transmission populations such as long-term care facilities and hospitals should be considered. These data also support the ongoing effort to develop second generation vaccines that might prevent acquisition of SARS-CoV-2.

## Data Availability

As the trial is ongoing, access to patient-level data and supporting clinical documents with qualified external researchers may be available upon request and subject to review once the trial is complete.

## Declaration of interests

The following authors have nothing to declare: AT, JA, AKR, LB, NE, NG LC reports grant funding from the NIAID/NIH.

## Acknowledgements

We thank the participants in the trials and the members of the Ubuntu and Sisonke trial teams for their dedication and contributions to the trial. We wish to thank Peter Meewes and Gillian Hunt for running assays, and Dr. Mindy Miner for technical editing.

Funding was provided by NIAID/NIH UM1 AI068614-14 (CoVPN LOC), UM1 AI068635 (CoVPN SDMC); 3UM1AI068618-15S1 (CoVPN LC). The Sisonke study was funded by the National Treasury of South Africa, the National Department of Health, Solidarity Response Fund NPC, The Michael & Susan Dell Foundation, The Elma Vaccines and Immunization Foundation - Grant number 21-V0001, and the Bill & Melinda Gates Foundation – grant number INV-030342.

The funders had no role in data collection and analysis, decision to publish, or preparation of the manuscript.

The study was approved by the South African Health Products Regulatory Authority (SAHPRA); Human Research Ethics Committee (Medical), University of the Witwatersrand, Johannesburg, South Africa; and

Human Research Ethics Committee, Faculty of Health Sciences, University of Cape Town, Cape Town, South Africa. All necessary patient/participant consent has been obtained and the appropriate institutional forms have been archived.

## Notes

### Competing Interest Statement

LC reports grant funding from the NIAID/NIH. All other authors have nothing to declare

### Clinical Trial

PACTR202105817814362

### Author Declarations

The study was approved by the South African Health Products Regulatory Authority (SAHPRA); Human Research Ethics Committee (Medical), University of the Witwatersrand, Johannesburg, South Africa; and Human Research Ethics Committee, Faculty of Health Sciences, University of Cape Town, Cape Town, South Africa. All necessary patient/participant consent has been obtained and the appropriate institutional forms have been archived.

### Summary of Updates

Authors and acknowledgements updated.

