## Supplemental material for "High Rate of Asymptomatic Carriage Associated with Variant Strain Omicron" (from another version of this preprint: version 1, 2021-12-27)

**Supplemental Table 1a. SARS-CoV-2 PCR positivity by SARS-CoV-2 serostatus and CD4+ cell count in PLWH.**

| | **Omicron detected** | **Omicron not detected** | **Total** | **% Omicron detected** |
|---|---|---|---|---|
| **Group 1** (SARS-CoV-2 seronegative) | | | | |
| 500 and above | 10 | 48 | 58 | 17% |
| 200 - 500 | 16 | 19 | 35 | 46% |
| less than 200 | 1 | 6 | 7 | 14% |
| Total | 27 | 73 | 100 | 27% |
| **Group 2** (SARS-CoV-2 seropositive) | | | | |
| 500 and above | 12 | 31 | 43 | 28% |
| 200 - 500 | 3 | 12 | 15 | 20% |
| less than 200 | 1 | 1 | 2 | 50% |
| Total | 16 | 44 | 60 | 27% |
| **Total** | **43** | **117** | **160** | **27%** |

**Supplemental Table 1b: Summary of cycle threshold (CT) values by Fisher Taqpath for those samples with S gene drop out (n=56).**

| **Target** | **N** | **Mean CT** | **SD CT** | **Min CT** | **Median CT** | **Max CT** |
|---|---|---|---|---|---|---|
| N gene | 56 | 25·8 | 5·3 | 14·5 | 25·8 | 35·7 |
| ORF1ab | 56 | 25·2 | 5·5 | 14·4 | 25·7 | 34·1 |
| Average N and ORF1a | 56 | 25·4 | 5·4 | 14·4 | 25·8 | 34·9 |

Note CT values of 20 and 25 correspond to log10 viral loads of approximately 6.5 and 5, respectively.

**Supplemental Table 2. Prevalence of asymptomatic SARS-CoV-2 infection by PCR in Sisonke sub study from enrollment through 6 months.**

| | **Predominant variant(s)*** | **No. of participants** | **SARS-CoV-2 PCR+** | **Prevalence** |
|---|---|---|---|---|
| **Enrollment** | Beta, Delta | 1,604 | 39 | 2·4% |
| **Week 3** | Delta | 1,447 | 51 | 3·5% |
| **Week 6** | Delta | 1,260 | 25 | 2·0% |
| **Month 6** | Omicron | 577 | 91 | 16% |
| ***HIV positive*** | *Omicron* | *169* | *27* | *16%* |
| ***HIV negative*** | *Omicron* | *405* | *62* | *15·3%* |

*Source: https://covariants.org/per-country)

Month 6 participants sub divided by HIV status are in italics.

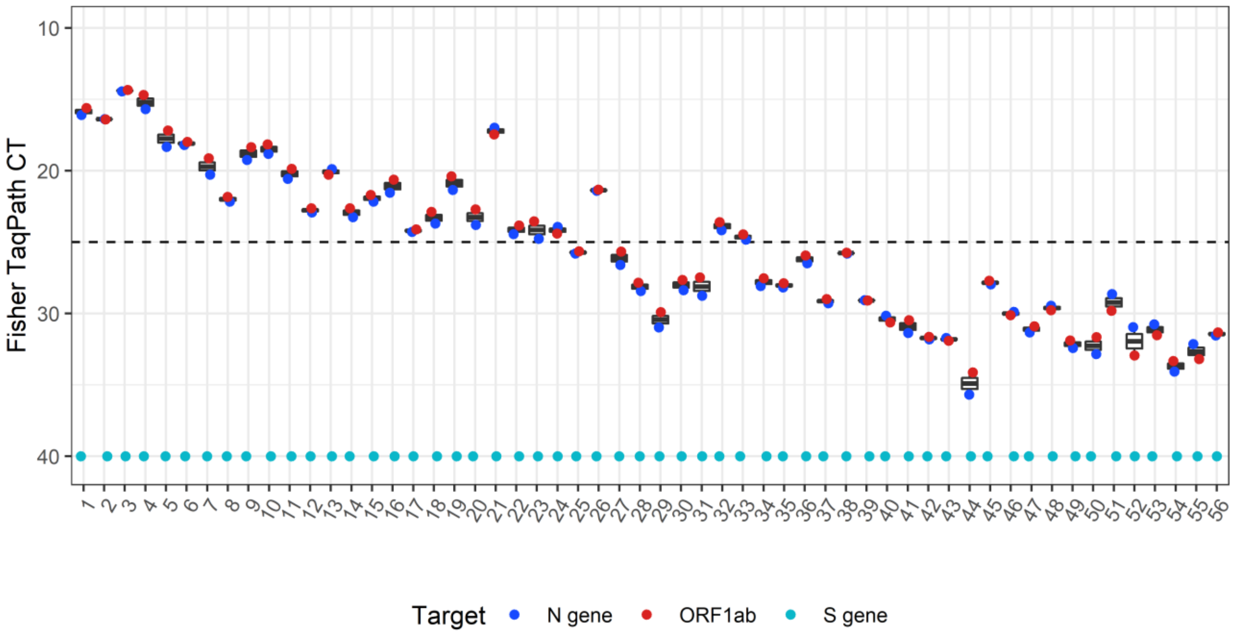


**Supplemental Figure 1. Summaries of cycle threshold values.** CoVPN 3008 baseline positive swabs (x axis) with S gene target failure (n=56). Fisher Taqpath CTs arranged in order of increasing Abbott cycle number. Dotted line denotes CT of 25. Note CT values of 20 and 25 correspond to log10 viral loads of approximately 6.5 and 5, respectively.
